# Evaluating a Medical-Grade Voice AI for Patient and Caregiver Guidance: A Multi-Scenario Nurse Panel Study

**DOI:** 10.1101/2025.09.18.25336107

**Authors:** Sahitya Sridhar, Rajashekar Vasantha, Supreet Deshpande, Parul Pathak

**Author notes:** **Corresponding Author** Sahitya Sridhar SynthioLabs San Francisco, CA, USA.

## Abstract

Medical-grade conversational AI offers the potential to extend patient support programs (PSPs) in regulated therapeutic areas, but its safety and reliability must be rigorously evaluated. We conducted a large-scale, nurse-led assessment of a voice-based AI system across 30 patient and caregiver scenarios spanning diabetes, oncology, neurology, cardiometabolic disease, and rare disorders. Nearly 1,000 U.S.-licensed nurses role-played patients or caregivers in more than 20,000 interactions, scoring the AI across five domains: clinical accuracy, empathy, communication clarity, appropriateness of advice, and compliance with evidence. The system achieved over 97% top ratings across all domains, with empathy noted most strongly in oncology and rare disease caregiving contexts, and clarity reflecting minor opportunities in pacing and call-closing behavior. Qualitative feedback emphasized tone, personalization, and regulatory compliance as consistent strengths. These findings demonstrate that voice-based AI can safely and effectively support patient and caregiver interactions, suggesting readiness for scaled deployment in pharma-led PSPs to expand after-hours coverage, provide consistent patient engagement, and generate feedback loops to inform both AI and human nurse training.

## 1. Introduction

Digital health interventions are increasingly used to support patients outside of traditional clinical encounters. In particular, conversational AI offers the possibility of scaling engagement while maintaining clinical-grade safety and empathy. However, regulatory concerns around accuracy, off-label avoidance, and patient trust have limited adoption in high-stakes settings such as patient support programs (PSPs).

This study aimed to rigorously evaluate a medical-grade voice AI system developed for life sciences applications. We designed a large-scale, nurse-led panel assessment to measure its performance in real-world therapeutic contexts, focusing on safety-critical domains such as accuracy, empathy, clarity, appropriateness of advice, and risk of inaccurate claims.

## 2. Methods

### 2.1 Study Design

The evaluation involved 30 patient and caregiver scenarios spanning oncology, cardiometabolic, neurology, dermatology, and rare disease conditions. Scenarios were derived from common PSP interactions, including missed doses, cold-chain travel, pediatric administration, and caregiver stress management.

### 2.2 Participants

A total of ∼1,000 U.S.-licensed nurses participated. Nurses were recruited across diverse states and specialties, representing 20+ years of average clinical experience. Each nurse was randomly assigned scenarios to role-play as patients or caregivers.

### 2.3 Procedure

- **Briefing:** Nurses received standardized role-play instructions for each scenario.
- **Interaction:** Each interacted live with the AI voice system in real time.
- **Evaluation:** After the interaction, nurses scored the AI across five domains: clinical accuracy, empathy, communication clarity, appropriateness of advice, and risk of inaccurate claims.
- **Qualitative Feedback:** Free-text notes were collected to capture narrative impressions and improvement suggestions.
- **Compliance Checks:** Nurses flagged any off-label promotion, lack of escalation, or inappropriate safety language.

### 2.4 Measures

- **Clinical Accuracy:** Concordance with product labeling and standard of care.
- **Empathy:** Validating patient emotions, offering reassurance.
- **Communication Clarity:** Use of plain language and logical sequencing.
- **Appropriateness of Advice:** Staying within the AI’s informational role, escalating when necessary.
- **Compliance with Evidence:** Avoiding overstatements or unfounded certainty.

### 2.5 Feedback-to-Improvement Cycle

To ensure continuous refinement of the AI system, we implemented a structured six-step loop: (1) scenario role-play by licensed nurses, (2) evaluator feedback, (3) challenge identification, (4) model refinement, (5) retesting and validation, and (6) performance uplift. This cyclical process (Figure 1) supported rapid iteration while maintaining compliance and safety standards.

**Figure 1.**
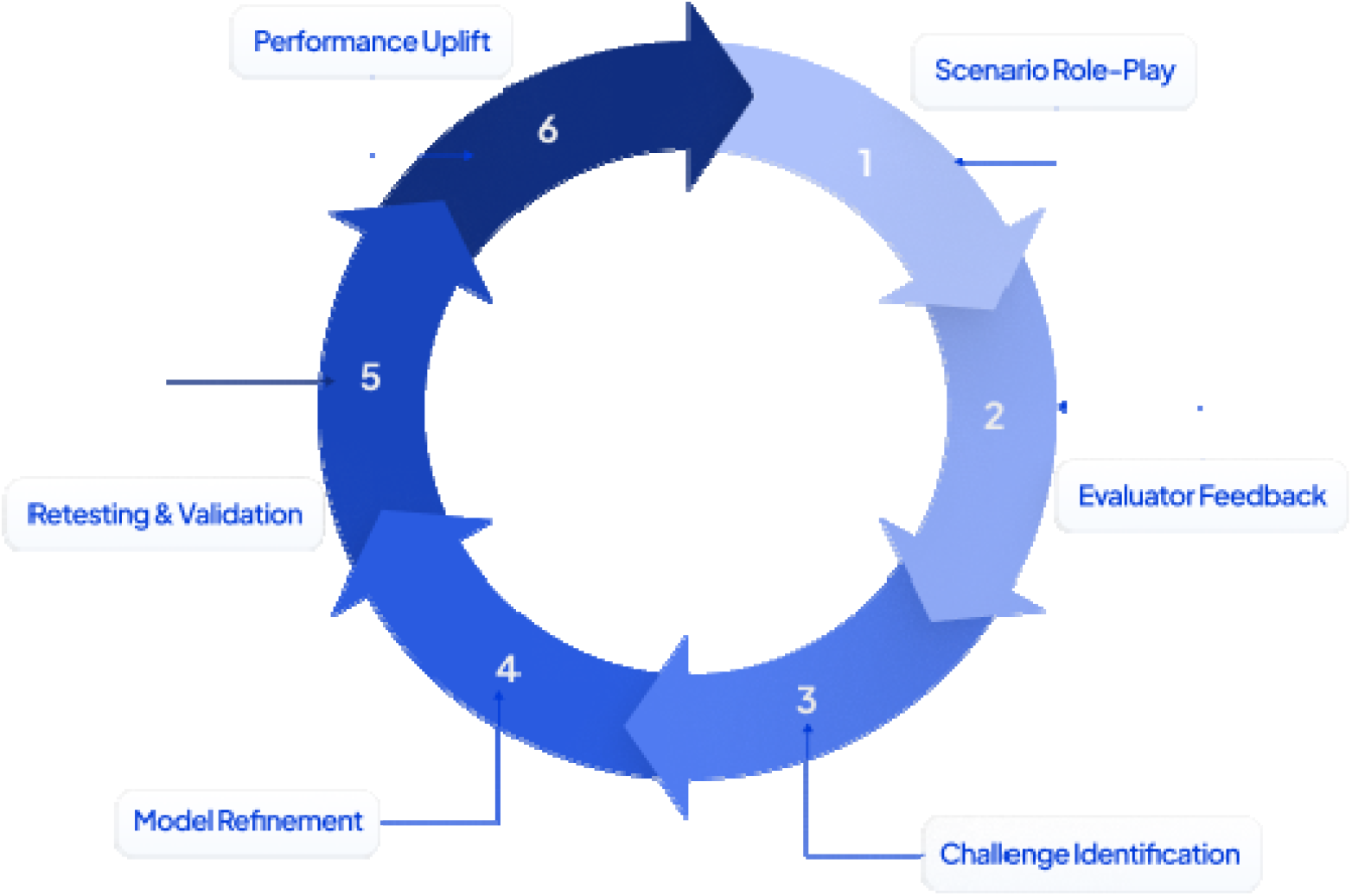
Feedback-to-Improvement Cycle. A six-step cycle showing how nurse role-play evaluations fed into model refinement. Steps included (1) Scenario Role-Play, (2) Evaluator Feedback, (3) Challenge Identification, (4) Model Refinement, (5) Retesting & Validation, and (6) Performance Uplift.

## 3. Results

### 3.1 Quantitative Outcomes

Nurses rated each interaction across five domains using a **three-point scale**:

- **Top (1):** Full demonstration of the domain (e.g., clinically accurate, empathetic, or clear with no gaps).
- **Mid (2):** Acceptable performance with minor limitations (e.g., slightly rushed pacing, somewhat abrupt escalation).
- **Low (3):** Missed expectations, requiring correction (e.g., unclear instruction, missed escalation opportunity).

Across 20,000+ evaluated interactions:

| Domain | Top (1) | Mid (2) | Low (3) |
| --- | --- | --- | --- |
| Clinical Accuracy | 97.8% — “Always matched product labeling, never strayed off-label.” | 2.2% | 0% |
| Appropriateness of Advice | 97.1% — “Consistently set boundaries and escalated when needed.” | 2.6% | 0.3% |
| Compliance with Evidence | 96.8% — “Careful with unknowns; avoided overstating.” | 3.2% | 0% |
| Empathy | 98% — *“Felt calm, reassuring, almost like talking to a real nurse.”* | 2% | 0% |
| Communication Clarity | 99% — *“Plain language, explained step by step.”* | 1% | 0% |

### 3.2 Domain Trends

- **Accuracy & Advice:** Strongest performance across all therapeutic clusters, especially in dosing and adherence contexts.
- **Empathy:** Widely praised in oncology and rare disease caregiving scenarios.
- **Clarity:** Slightly lower scores linked to rapid pacing and early call wrap-ups.

### 3.3 Qualitative Strengths

- Regulatory Compliance: “*No off-label promotion; consistent escalation when needed.”*
- Empathy: “*Tone was calm and reassuring.”*
- Contextual Adaptation: “*It adjusted well to caregiver vs. patient personas.”*
- Safety Language: “*Clear framing of knowns vs. unknowns.”*

### 3.4 Areas for Refinement

- Pacing: Nurses noted occasional rapid delivery during complex instructions.
- Closing Behavior: Calls sometimes ended before confirming additional patient questions.

## 4. Discussion

This study provides large-scale evidence that voice-based AI can safely and effectively support patient and caregiver interactions across diverse therapeutic areas. The high top-box scores (>97% across all domains) demonstrate consistent clinical accuracy and empathy — critical dimensions for trust in regulated healthcare contexts.

Notably, qualitative nurse feedback highlighted the AI’s ability to adapt communication style depending on persona (e.g., slower and calmer with grandparents, concise with teenagers). This adaptability suggests potential for personalization at scale.

Opportunities for refinement include improving pacing during procedural guidance and reinforcing structured call-closing prompts. However, these areas represented <3% of total feedback and did not undermine overall performance.

## Data Availability

All data produced in the present study are available upon reasonable request and will be considered on a case-by-case basis by the corresponding authors.

## 5. Implications for PSPs

The findings of this study suggest several practical applications for pharmaceutical patient support programs (PSPs). **Scalability** emerged as a core strength: the AI was able to handle a high volume of simulated patient interactions without loss of accuracy, pointing toward its potential as a force multiplier for nurse call centers. **After-hours coverage** is another key implication; by providing consistent, empathetic guidance outside of traditional nurse availability, the system can reduce patient anxiety and ensure continuity of support. The AI also functions as a **training tool**, with aggregated nurse feedback highlighting opportunities to refine human PSP scripts and educational content.

Finally, the system demonstrated strong **regulatory readiness**, consistently aligning responses with approved product labeling and escalation standards — an essential requirement for deployment in life sciences contexts.

## 6. Future Work

Future development of the system will focus on extending both accessibility and personalization. A **multilingual rollout** is planned, beginning with Spanish, French, and Mandarin, to address diverse patient populations and reduce language barriers in PSP delivery. Enhancements in **behavioral personalization** will allow the AI to adapt tone, pacing, and level of detail to the patient’s communication style, thereby improving trust and comprehension. Finally, **deeper metrics** are needed to complement evaluator scoring, including structured post-call comprehension testing to assess how well patients retain information provided during AI interactions. Together, these directions point toward a more adaptive and inclusive AI system that can expand the reach and impact of patient support services.

## Notes

### Competing Interest Statement

The authors are affiliated with SynthioLabs, the company that developed the evaluated AI system. No other competing interests declared.

### Funding Statement

This study was fully funded by SynthioLabs. No external funding, grants, or third-party services were received for the design, conduct, analysis, or reporting of this work.

